# ABERRANT IMMUNE PROGRAMMING IN NEUTROPHILS IN CYSTIC FIBROSIS

**DOI:** 10.1101/2023.01.22.23284619

**Authors:** Yawen Hu, Christine M. Bojanowski, Clemente J. Britto, Dianne Wellems, Kejing Song, Callie Scull, Scott Jennings, Jianxiong Li, Jay K. Kolls, Guoshun Wang

## Abstract

Cystic fibrosis (CF) is a life-shortening genetic disorder, caused by mutations in the gene that encodes Cystic Fibrosis Transmembrane-conductance Regulator (CFTR), a cAMP-activated chloride and bicarbonate channel. Although multiple organ systems can be affected, CF lung disease claims the most morbidity and mortality due to chronic bacterial infection, persistent neutrophilic inflammation, and mucopurulent airway obstruction. Despite the clear predominance of neutrophils in these pathologies, how CFTR loss-of-function affects these cells *per se* remains incompletely understood. Here, we report the profiling and comparing of transcriptional signatures of peripheral blood neutrophils from CF participants and healthy human controls (HC) at the single-cell level. Circulating CF neutrophils had an aberrant basal state with significantly higher scores for activation, chemotaxis, immune signaling, and pattern recognition, suggesting that CF neutrophils in blood are prematurely primed. Such an abnormal basal state was also observed in neutrophils derived from an F508del-CF HL-60 cell line, indicating an innate characteristic of the phenotype. LPS stimulation drastically shifted the transcriptional landscape of HC circulating neutrophils towards a robust immune response, however, CF neutrophils were immune-exhausted. Moreover, CF blood neutrophils differed significantly from CF sputum neutrophils in gene programming with respect to neutrophil activation and aging, as well as inflammatory signaling, highlighting additional environmental influences on the neutrophils in CF lungs. Taken together, loss of CFTR function has intrinsic effects on neutrophil immune programming that leads to premature priming and dysregulated response to challenge.

## INTRODUCTION

Cystic fibrosis (CF) is a chronic, progressive and debilitating genetic disorder, caused by mutations in the Cystic Fibrosis Transmembrane-conductance Regulator gene (*CFTR*) that encodes a cAMP-activated chloride and bicarbonate channel (1, 2). Clinically, CF affects multiple organ systems, including the lung, intestines, pancreas, liver, and reproductive system. However, the lung disease claims the most morbidity and mortality. Dominant pathological changes in the lung are chronic bacterial infection, persistent neutrophilic inflammation, and mucopurulent airway obstruction (3, 4). Highly effective CFTR modulator therapies can effectively correct CF epithelial defects, reflected by normalization of the sweat Cl^−^ transport (5, 6). However, the neutrophilic inflammation persists, suggesting a potentially different pathogenic mechanism that the current modulators are not known to target.

Polymorphonuclear neutrophils (PMN) are a major cellular component in host defense against extracellular bacterial infection, and are the first line of immune cells recruited to sites of infection and inflammation (7). The principal function of PMN is to contain, degrade and eradicate invading microorganisms through phagocytosis, cytotoxicity, and enzymatic digestion (8, 9). Once a microbial challenge is addressed, PMN are cleared by macrophages through efferocytosis to restore tissue homeostasis (10). CF lungs exhibit unrelenting neutrophilic inflammation, suggesting an abnormal immune response. However, how CF neutrophils directly contribute to this pathogenesis is not clearly known.

Single-cell RNA sequencing (scRNA-seq) is a powerful tool to infer cell behavior and function through capture and deciphering of the transcriptional dynamics of individual live cells. Previous publications have elegantly used this method to study human neutrophil heterogeneity (11), and transcriptional dynamics at different states (12). Here, we performed scRNA-seq on peripheral blood neutrophils from CF participants and healthy human controls (HC) to profile and compare their molecular signatures at basal state or upon challenge. CF circulating PMN were also compared to CF sputum PMN to understand transcriptional reprogramming after lung recruitment. Our work has demonstrated that CFTR loss-of-function in neutrophils alters the cell intrinsic immune programming that is further influenced by the CF lung environment. This transcriptional defect results in neutrophil premature priming and dysregulated immune response to challenge.

## RESULTS

### Circulating neutrophils from CF blood are abnormally primed and dysregulated in response to LPS challenge

Peripheral blood neutrophils from CF and HC subjects were isolated by Percoll density gradient centrifugation (13). All the CF participants were F508del homozygotes receiving Elexacaftor/Tezacaftor/Ivacaftor modulator therapy and with stable disease. Human subject demographics are provided in **Supplementary Table 1**. The obtained neutrophils were divided into two aliquots: one for immediate processing for scRNA-seq, and the other for culture and exposure to *Pseudomonas aeruginosa* LPS for 16 hours, followed by scRNA-seq. Cell viability of each condition was 90% or greater. As the cells were purified largely to a pure neutrophil population, after rigorous quality control (**Supplementary Figs. 1-4**), we readily obtained 26,568 high-quality cells with a dataset composed of 4 experimental conditions, including 6,563 HC neutrophils (Neutrophils from health controls without any culture or treatment), 5,600 HC_LPS neutrophils (Neutrophils from health controls with culture and LPS challenge), 7,698 CF neutrophils (Neutrophils from CF participants without any culture or treatment), and 6,707 CF_LPS neutrophils (Neutrophils from CF participants with culture and LPS challenge). Uniform Manifold Approximation and Projection (UMAP) was computed of the 4 groups of cells. As shown (**Fig. 1a**), HC and CF neutrophils had an overlapping UMAP distribution, while LPS challenge shifted the projection pattern of the cells. Separate plotting according to genotype provided a discrete view of UMAP for each genotype (**Fig. 1b & c**).

**Fig. 1.**
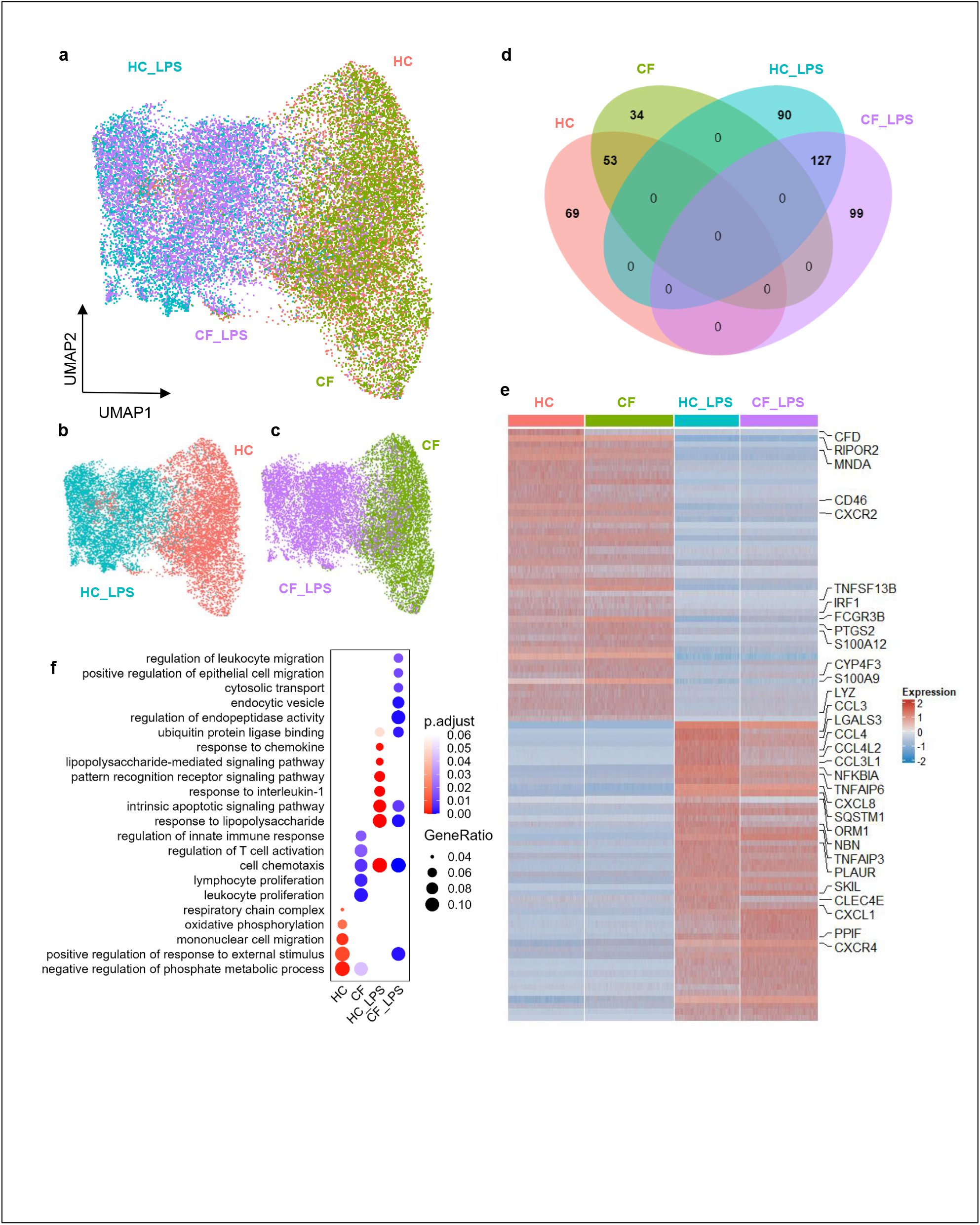
Atlas and transcriptional signatures of peripheral blood neutrophils from CF patients and healthy controls (HC). **a**, UMAP of peripheral blood neutrophils from their basal state and LPS-stimulated activating state. There are 4 groups of cells: 1) HC, colored in red, 2) HC_LPS, colored in blue, 3) CF, colored in green, and 4) CF_LPS, colored in purple. **b**, UMAP of neutrophils from HC and HC_LPS groups. **c**, UMAP of neutrophils from CF and CF_LPS groups. **d**, Venn diagram showing similarities of DEGs among the 4 groups of neutrophils, **e**, Heatmap showing the top 30 DEGs from each group. Selected immune related genes are marked. **f**, Dotplot showing Gene Ontology (GO) enrichment of DEGs from 4 groups.

Similarities of differentially expressed genes (DEGs, Log_2_FC>0.25, p<0.05) among the 4 groups were analyzed and are displayed in the Venn diagram (**Fig. 1d**). HC and CF neutrophils shared a significant number of DEGs (53 of 122 for HC and 53 of 87 for CF). Strikingly, LPS stimulation drastically changed the gene programming in both CF and HC neutrophils, reflected by zero DEGs shared between HC and HC_LPS and between CF and CF_LPS. Comparison between HC_LPS and CF_LPS found 127 shared items (Log_2_FC>0.25, p<0.05) (**Fig. 1d**). The top 30 DEGs from each group were profiled with immune-related genes marked (**Fig. 1e**). LPS stimulation activated a different set of top DEGs (**Fig. 1e**). Gene Ontology (GO) analysis revealed the top 5 predominant GO terms of each group (**Fig. 1f**). HC neutrophils showed enrichment of the gene transcripts related to Negative regulation of phosphate metabolic process (GO:0045936), Mononuclear cell migration (GO:0071674), Response to external stimulus (GO:0032103), Respiratory chain complex (GO:0098803), and Oxidative phosphorylation (GO:0006119), representing the gene signature of healthy circulating neutrophils at their basal state. In contrast, neutrophils from the CF participants showed enrichment of the gene transcripts related to Leukocyte proliferation (GO:0070661), Lymphocyte proliferation (GO:0046651), Cell chemotaxis (GO:0060326), Regulation of innate immune response (GO:0045088), and Regulation of T cell activation (GO:0050863), suggesting a status of pro-inflammatory priming. Furthermore, HC neutrophils exposed to LPS upregulated the typical gene clusters related to LPS response, such as Response to IL-1, Pattern recognition receptor signaling pathway, and LPS-mediated signaling pathway. However, CF_LPS neutrophils differed greatly from HC_LPS neutrophils with the most enhanced gene expressions related to ubiquitin protein ligase binding, regulation of endopeptidase activity, and endocytic vesicle formation, indicating an aberrant response to LPS (**Fig. 1f**).

To confirm the above findings, gene scores for immune-related properties or functions were quantitatively assessed using the standard GO terms. Measurement of neutrophil activation, based on 21 signature genes (*ACKR2, ANXA3, CCL5, CXCL8, CXCR2, CXCR4, F2RL1, FCER1G, KMT2E, MYO1F, PREX1, PRG3, PRKCD, STX11, STXBP2, STXBP3, SYK, TYROBP, VAMP2, VAMP7*, and *VAMP8*) (GO: 0042119), indicated that CF neutrophils in circulation had a significantly higher activation score than HC neutrophils, verifying that these cells in their basal state had been primed for pro-inflammatory response. Surprisingly, under LPS stimulation CF blood neutrophils had a significantly lower activation score than HC neutrophils (**Fig. 2a**), indicating a blunted immune response. These findings were further confirmed by significantly higher scores of CF neutrophils for Chemotaxis (GO:0030593) and Cytokine-related signaling (GO:0019221) at their basal state in circulation, but significantly lower scores of these immune properties upon LPS challenge (**Fig. 2b & c**). Moreover, CF neutrophils also had significantly lower scores for Neutrophil migration (GO:1990266) and Immune response-regulating signaling (GO:0002764) under LPS stimulation (**Fig. 2d & e**). Furthermore, the scores of specific signaling pathways that are highly related to neutrophil innate immune functions were also calculated. Toll-like receptor signaling (GO:0002224), Pattern recognition receptor signaling (GO:0002221), and LPS response signaling (GO:0032496) in CF circulating neutrophils had significantly higher scores than those in HC circulating neutrophils at their basal state (**Fig. 2f - h**), but with LPS challenge these signaling pathways were significantly down-regulated (**Fig. 2f - h**). All these data support the conclusion that CF blood neutrophils are abnormally primed at their basal state, and subdued in response to challenge.

**Fig. 2.**
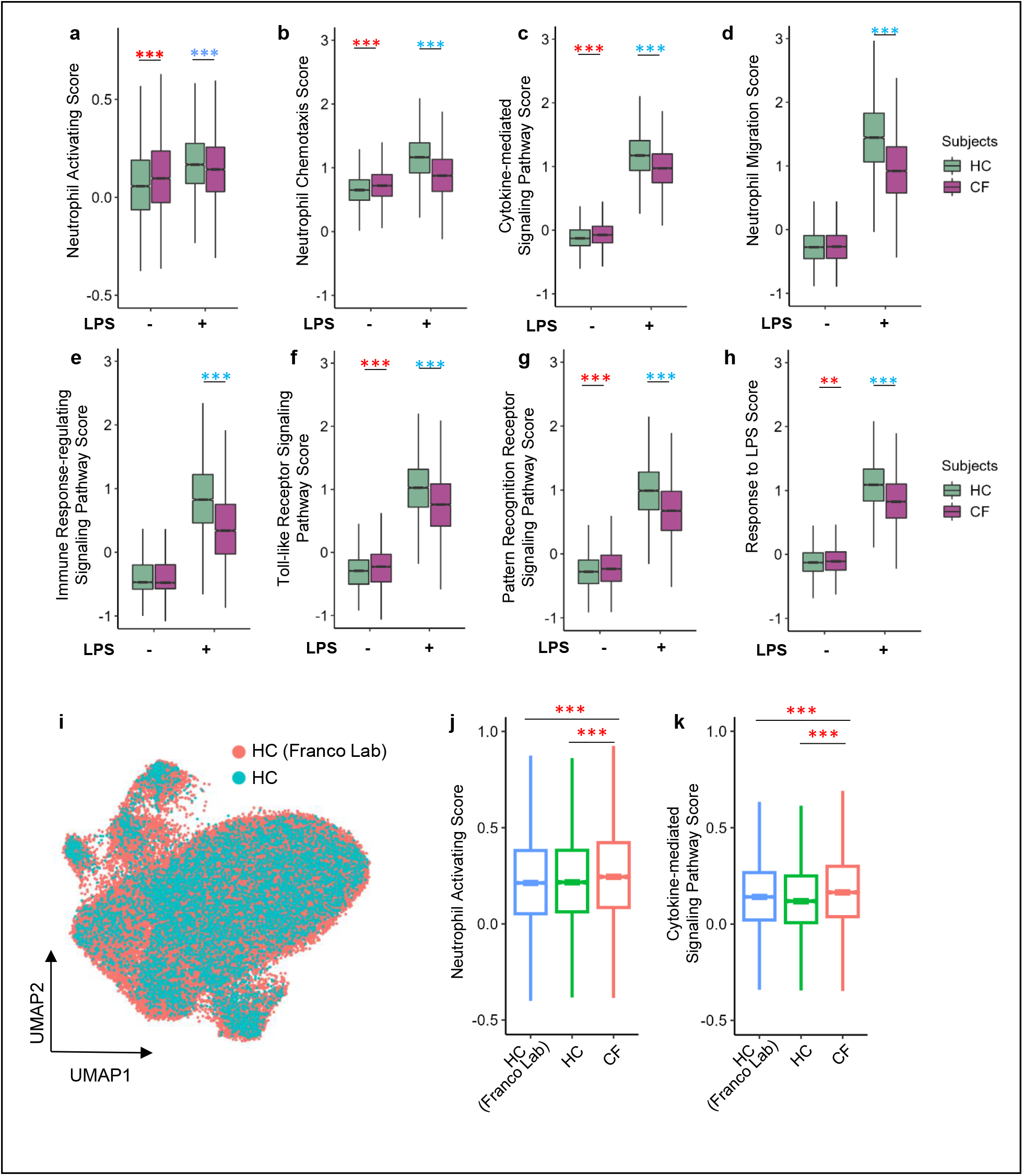
Gene scores for immune properties of CF and HC neutrophils at resting and upon LPS stimulation. **a-h**, Boxplots showing neutrophil immune scores of HC and CF neutrophils in resting (– LPS) and LPS stimulated (+ LPS) conditions. **a**, Neutrophil activation score (GO: 0042119). **b**, Neutrophil chemotaxis score (GO: 0030593). **c**, Score for cytokine-mediated signaling pathway (GO: 0019221). **d**, Score for neutrophil migration (GO: 1990266). **e**, Score for immune response-regulating signaling pathway (GO: 0002764). **f**, Score for toll-like receptor signaling pathway (GO: 0002224). **g**, Score for pattern recognition receptor signaling pathway (GO: 0002221). **h**, Score for response to LPS (GO: 0032496). **i**, UMAP showing dataset integration of our HC cohorts and selected Franco Lab HC cohorts. **j & k**, Box plots showing two immune function scores in a 3-group comparison. **j**, Neutrophil activation score (GO: 0042119). **k**, Score for cytokine-mediated signaling pathway (GO: 0019221). ANOVA test was used to judge statistical significance between two comparing groups. *p<0.05, **p<0.01, ***p<0.001. Red asterisks indicate that CF neutrophils have significantly higher score and blue asterisks significantly lower score in each comparison with HC counterparts.

To rule out the possibility that the observed transcriptomic difference between HC and CF neutrophils was derived from different cell isolation methods or subject disparities, we compared our dataset with that of selected cohorts from the published dataset by Franco Lab (14), for which their neutrophils were isolated by negative selection (**Supplementary Table 1**). Results indicated that the 2 HC datasets integrated well in a good overlapping UMAP (**Fig. 2i**). Three-group comparisons demonstrated that the neutrophil activation score and cytokine-mediated signaling pathway score of the CF circulating neutrophils were significantly higher than those of Franco Lab HC cohorts, validating the results from comparing with our HC cohorts (**Fig. 2 j & k**). Thus, circulating neutrophils in CF blood indeed have an aberrant immune transcriptional program at their basal state.

### CF and healthy neutrophils show differential state transition upon LPS challenge

Normal neutrophils upon challenge undergo a programmed transformation from basal state to activating state, and further to aging state. Monocle trajectory of our HC data revealed such three states (**Fig. 3a**), which correlated well with experimental conditions (with or without LPS) (**Fig. 3b**). Specifically, basal state HC cells were from the non-LPS group, while the cells in activating and aging states were predominantly from the LPS-treated group. Intriguingly, LPS stimulation resulted in two distinct subpopulations, showing a bifurcation in trajectory (**Fig. 3b**). Cells at the three states from the HC cohorts were color-mapped in UMAP (**Fig. 3c**). GO analysis was conducted to define the gene signature of each state. The basal state HC neutrophils highly enriched the transcripts related to Actin filament organization, Negative regulation of phosphate metabolic process, and Leukocyte cell adhesion (**Fig. 3d**). Despite the fact that the activating state and aging state were all derived from LPS stimulation, they differed in their top GO enrichment. The gene clusters that were uniquely or significantly enriched at activating state were highly immune-related, and included Chemokine-mediated signaling, Proton transmembrane transport, Granulocyte migration, and Wound healing. In contrast, the top GO terms for the HC neutrophils at aging state were related to Negative regulation of endopeptidase activity, Negative regulation of proteolysis, Positive regulation of cell adhesion, Regulation of inflammatory responses, and Intrinsic apoptotic signaling, suggesting that the cells were progressing from an immune-active state to an immune-aging state and towards programmed cell death. CF neutrophils were found to share the general pattern in trajectories (**Fig. 3e & f**) with the HC neutrophils (**Fig. 3a & b**). Color-mapped UMAP for each state of the CF cohorts (**Fig. 3g**) showed a similar projection pattern as the HC cohorts (**Fig. 3c**). Composition analyses of the HC_LPS and CF_LPS cells demonstrated that the majority (∼66%) of HC_LPS cells were at activating state, while the remaining ∼33% cells at aging state (**Fig. 3h)**. In contrast, CF_LPS population at activating state occupied ∼37%, while the majority (∼62%) were at aging state (**Fig. 3h**). This result suggests that the majority of CF_LPS neutrophils were already further along in the aging/cell death trajectory. Similarity comparisons of DEGs of HC_LPS and CF_LPS neutrophils at either activating state or aging state revealed over 50% of shared DEGs at each state. However, there were a significant number of DEGs that were not shared between genotypes (**Fig. 3i & j**).

**Fig. 3.**
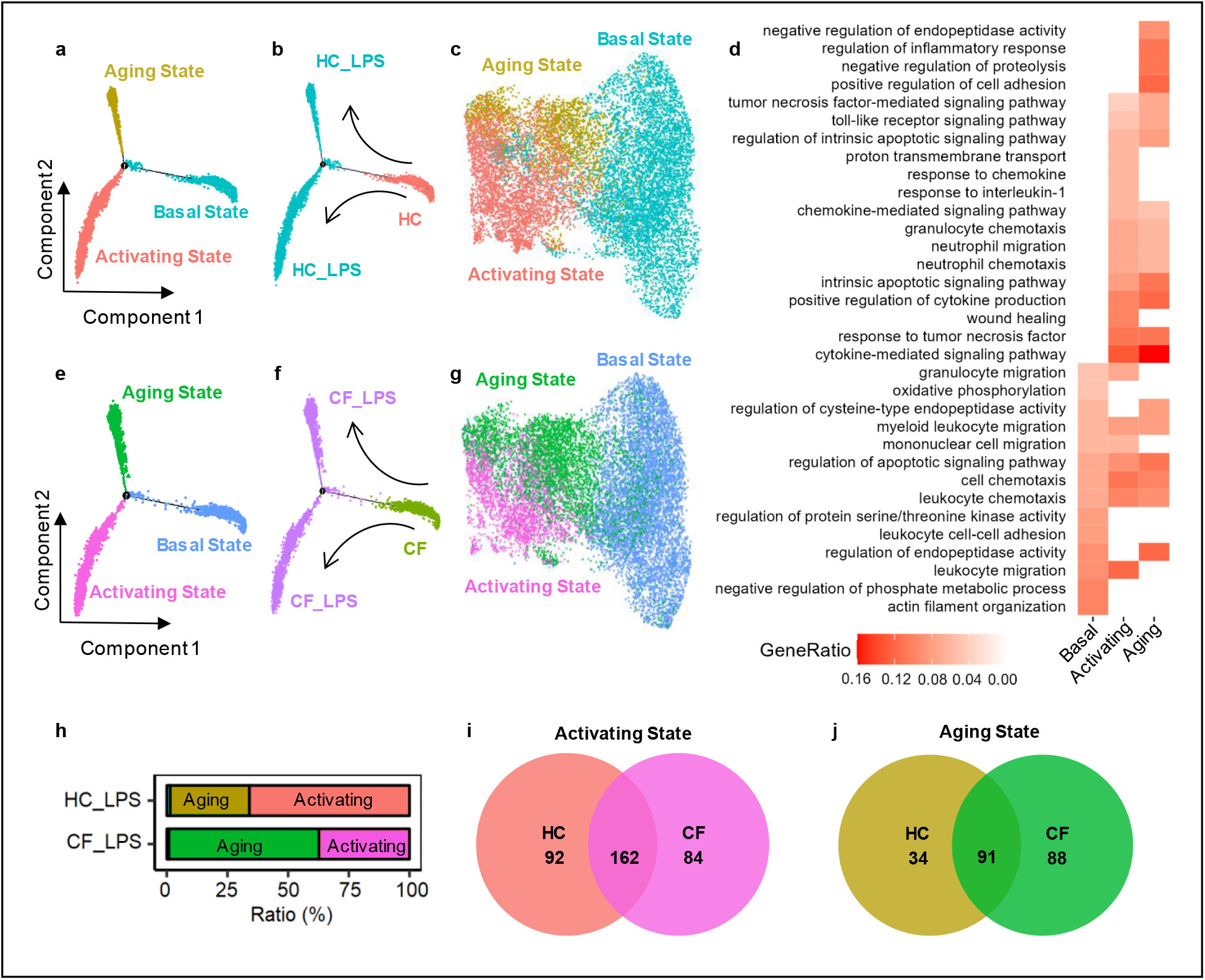
Cell state transition of CF and HC neutrophils in response to LPS. **a**, Monocle trajectory of HC neutrophils colored by states (Basal, Activating and Aging). **b**, Monocle trajectory of HC neutrophils colored by groups (HC and HC_LPS). **c**, UMAP of the neutrophils from HC and HC_LPS groups, colored by states. **d**, GO enrichment of DEGs in each state of HC neutrophils. **e**, Monocle trajectory of CF neutrophils, colored by states. **f**, Monocle trajectory of CF neutrophils, colored by groups. **g**, UMAP of the neutrophils from HC and HC_LPS groups, colored by states. **h**, Proportions of Activating state and Aging state in the population of HC_LPS or CF_LPS cells. **i & j**, Venn diagrams showing overlapping DEGs between HC neutrophils and CF neutrophils in the Activating state (**i**) or Aging state (**j**).

### CF and healthy neutrophils display differential cell-fate decision and cell-aging processes after LPS challenge

RNA velocity analysis is an effective method to predict cell-fate commitment and cell-aging processes using the information from newly transcribed pre-mRNAs (unspliced) and mature mRNAs (spliced) (15). Velocities derived from the dynamic models of HC and CF neutrophils were visualized as streamlines in their corresponding UMAP-based embedding, revealing the standing of each cell in their life journey and fate direction (**Fig. 4a & c**). Each arrow represented a velocity vector indicating the predicted direction and speed of progression along the direction of an individual cell. There were two starting points at the basal state for HC or CF, as circled (**Fig. 4a & c**), which correlated well with the root cells identified by terminal_states analysis (**Fig. 4b & d**). Upon LPS challenge, the majority of basal state HC neutrophils took the direction towards the aging state, the end point of their life journey (**Fig. 4c**). HC neutrophils in the activating state also pointed to the aging state. However, CF neutrophils had a strong velocity pattern that originated from the basal state, headed toward the aging state, and ended in the activating state (**Fig. 4c**). Such an unusual senile activation reflects an ill-selected fate and ill-programmed cell death, which could lead to a protracted inflammation.

**Fig. 4.**
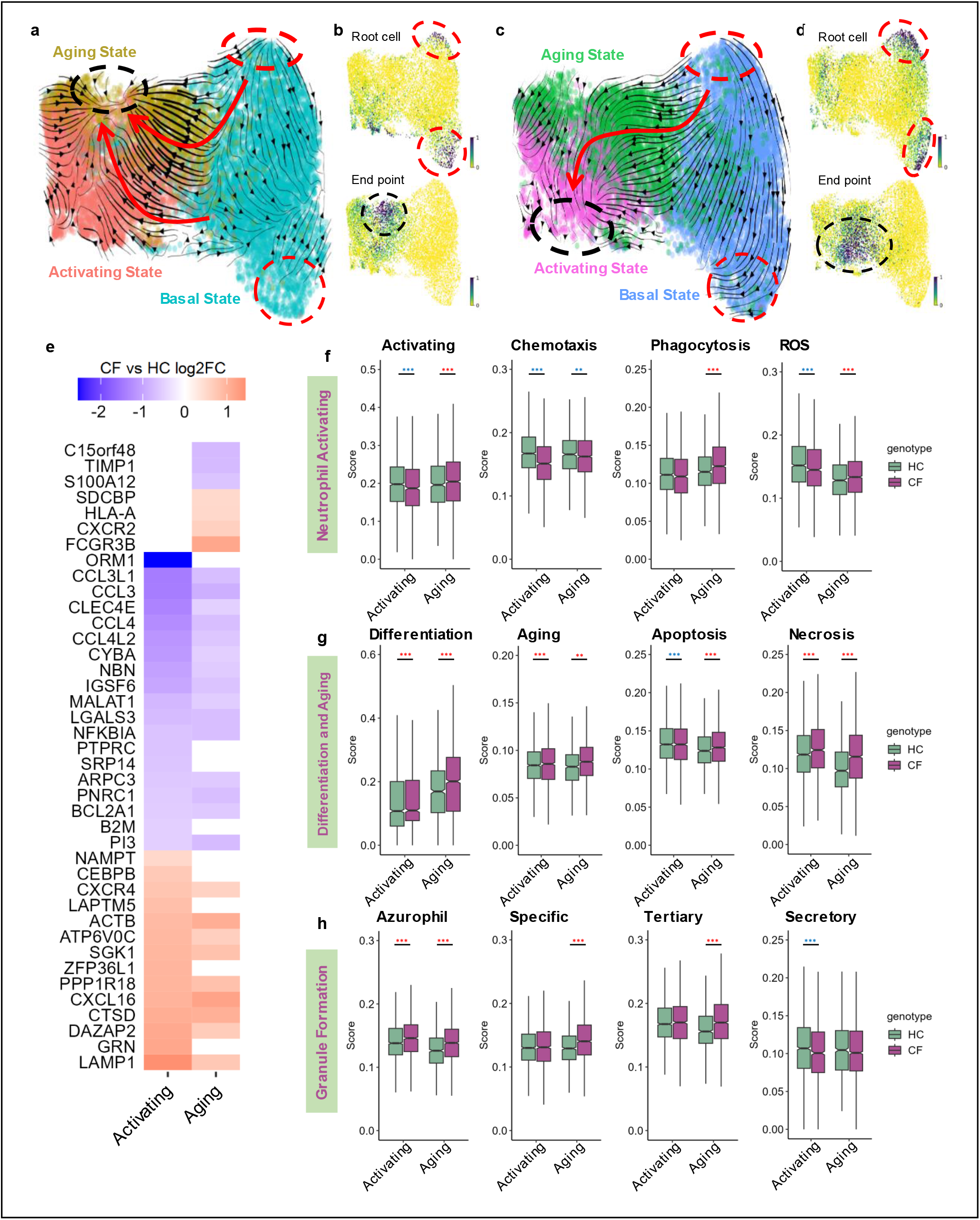
Cell fate decision and GO enrichment of CF and HC neutrophils in response to LPS. **a**, RNA velocity plot revealing the origin and inter-relationship of 3 states of HC neutrophils. **b**, UMAP-embedding root cells and end point of HC neutrophils. **c**, RNA velocity plot showing the origin and inter-relationship of 3 states of CF neutrophils. **d**, UMAP-embedding root cell and end point of CF neutrophils. **e**, Heatmap showing upregulated genes (red) and downregulated genes (blue) in CF samples in Activating state or Aging state, as compared to HC. **f-h**, Comparisons of neutrophil property and function scores between HC and CF neutrophils in Activating state or Aging state. **f**, Neutrophil activation scores; **g**, Differentiation and aging scores; **h**, Granule formation scores. Significance was determined by two-way ANOVA. *p<0.05, **p<0.01, ***p<0.001. Red asterisks indicate that CF neutrophils have significantly higher scores, and blue asterisks significantly lower scores, as compared to HC neutrophils.

To better define the gene signature of the activating or the aging states and to determine how CFTR mutation alters the gene enrichment in each state, the top 10 up-regulated and the top 10 down-regulated genes in CF neutrophils from comparison with the HC counterparts were inspected and are displayed in the heatmap (**Fig. 4e**). In the activating state, the most upregulated genes in CF neutrophils were related to regulating lysosomal functions (e.g. *GRN* and *LAPTM5*) and neutrophil development and survival (e.g. *NAMPT* and *CEBPB*), while the most down-regulated genes in CF neutrophils included acute phase protein gene (e.g. *ORM1*), CD45 coding gene *PTPRC*, and MHC I alpha chain coding gene *B2M* (**Fig. 4e**), which were associated with significantly lower scores for cell activation, chemotaxis, and reactive oxygen species (ROS) (**Fig. 4f**). Hence, while they had an early pro-inflammatory priming, the CF neutrophils upon challenge seemed to be immune-exhausted, reflected by significantly weakened immune activation and response. GO enrichment analysis also supported this conclusion by showing that CF neutrophils in the activating state enriched the transcripts related to Cell-substrate junction (GO:0030055), Focal adhesion (GO:0005925) and Epithelial cell migration (GO:0010631), which were not directly involved in neutrophil immune function (**Supplementary Fig. 5**). As for the aging state, the most upregulated genes in CF neutrophils included *FCGR3B, CXCR2, HLA-A*, and *SDCBP*, and the most downregulated genes where *S100A12, TIMP1*, and *C15orf48* (**Fig. 4e**). These regulations were associated with significantly higher scores for differentiation, aging, apoptosis, and necrosis, as compared to HC neutrophils in the same state (**Fig. 4g**). Because neutrophil immune function is highly associated with phagosomal function(16), we then examined the scores for granule formation (azurophil, specific, tertiary and secretory). These analyses showed that CF neutrophils in their activating state had a higher azurophil granule score but a lower secretory score, but in the aging state, the CF neutrophils had a significantly higher scores for azurophil, specific and tertiary granules (**Fig. 4h**). As granule proteins are normally pre-synthesized during neutrophil maturation(17), re-activation of these granule genes in the aging state confirmed a dysregulated gene programming for phagosomal function. These data indicate that CF neutrophils have irregular cell-fate decision and disordered cell activation and ending programs, which likely compromises their function and clearance.

### Neutrophils derived F508del-CF HL-60 cell line have a pro-inflammatory basal state and abnormal immune response to LPS

The identified pro-inflammatory basal state of CF circulating neutrophils can be determined by intrinsic neutrophil properties or induced by blood environmental factors. To explore this issue, we measured the levels of endotoxin and key pro-inflammatory cytokines (IL-8, IL-6, TNF-α and IL-1β) in the plasma samples of HC and CF subjects. All the measured samples had no detectable levels of the cytokines, and only one HC plasma had a very low level of endotoxin (∼0.07 U/ml) (**Supplementary Fig. 6**). These data led us to speculate that the aberrant transcriptional program in CF neutrophils is likely intrinsic. To further explore this possibility, we differentiated WT HL-60 cells (18) and F508del-CF HL-60 cells (19) with 1.25% DMSO into their corresponding neutrophil-like cells. ScRNA-seq was similarly performed and the gene signatures of these derived neutrophils with or without LPS stimulation were compared. There were 4 groups of samples: 1) WT_dHL-60 (WT neutrophils differentiated from wild-type HL-60 cell line), 2) CF_dHL-60 (CF neutrophils differentiated from F508del-CF HL-60 cell line), 3) WT_dHL-60_LPS (WT_dHL-60 stimulated with LPS), and 4) CF_dHL-60_LPS (CF_dHL-60 stimulated with LPS). UMAP for each genotype of cells was separately plotted, showing a comparable projection pattern (**Fig. 5a & b**). Top 5 GO enrichments indicated that WT_dHL-60 neutrophils largely enriched the transcripts related to immune cell development, while CF_dHL-60 cells most enriched those related to regulation of innate immune responses and granule formation (**Fig. 5c**), reflecting a pro-inflammatory priming. Upon LPS challenge, WT_dHL-60 neutrophils upregulated genes related to a typical immune response to LPS, such as Cytokine activity, Neutrophil chemotaxis, Neutrophil migration, Response to tumor necrosis factor, and LPS-mediated signaling pathway, while CF_dHL-60 cells failed in this response, and turned to antioxidant activity, collagen catabolic process, and antigen processing and presentation (**Fig. 5c**). Moreover, CF_dHL-60 neutrophils without LPS stimulation had a significantly higher neutrophil activation score (GO: 0042119) (**Fig. 5d**), and significantly upregulated the hallmark gene sets responsible for neutrophil immune functions (**Fig. 5h-k**). With LPS stimulation, CF_dHL-60 neutrophils had significantly lower scores for the immune features than WT_dHL-60 counterparts (**Fig. 5e-k**). These results corroborate the findings from the CF circulating neutrophils that CFTR mutation renders neutrophils pro-inflammatory in basal state and dysregulated in response to LPS challenge.

**Fig. 5.**
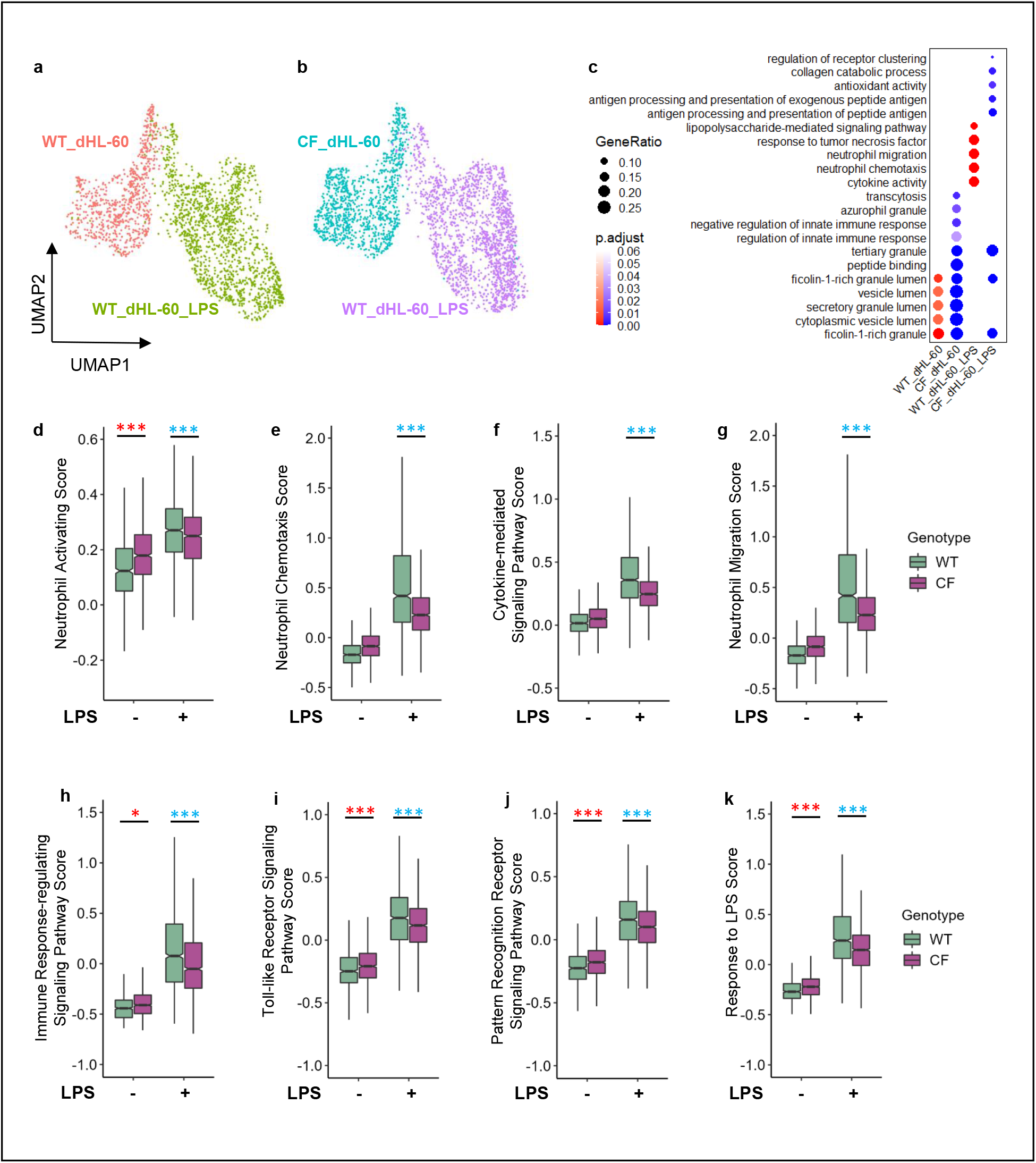
Pre-activation of neutrophils derived from CF HL-60 cells. **a**. UMAP of WT_differentiated HL-60 cells (WT_dHL-60). **b**. UMAP of CF_differentiated HL-60 cells (CF_dHL-60). **c**, Dot plot showing GO enrichment of DEGs in 4 groups. **d-k**, Box plots showing activation scores of neutrophils derived from WT and CF HL-60 cells with or without LPS stimulation. The significance was determined by two-way ANOVA. *p<0.05, **p<0.01, ***p<0.001. Red asterisks indicate that the derived CF neutrophils have significantly higher scores, and blue asterisks significantly lower scores, as compared to the derived WT neutrophils.

### CF lung environment further affects CF neutrophil gene programming

CF neutrophils mobilized to CF lungs have to go through extravasation, migration, adaptation, activation, cell death and clearance. To discern any differences in gene programming between circulating and airway neutrophils in CF, we compared our CF blood neutrophil dataset with a published one of CF sputum neutrophils (20). To match our CF patients’ demographic characteristics and treatments, only homozygous F508del CF subjects under similar modulator therapy were selected (**Supplementary Table 1**). Integration of the datasets gave rise to an overall UMAP showing 3 distinct cell congregations with unique DEG profiles (**Fig. 6a & b**).

**Fig. 6.**
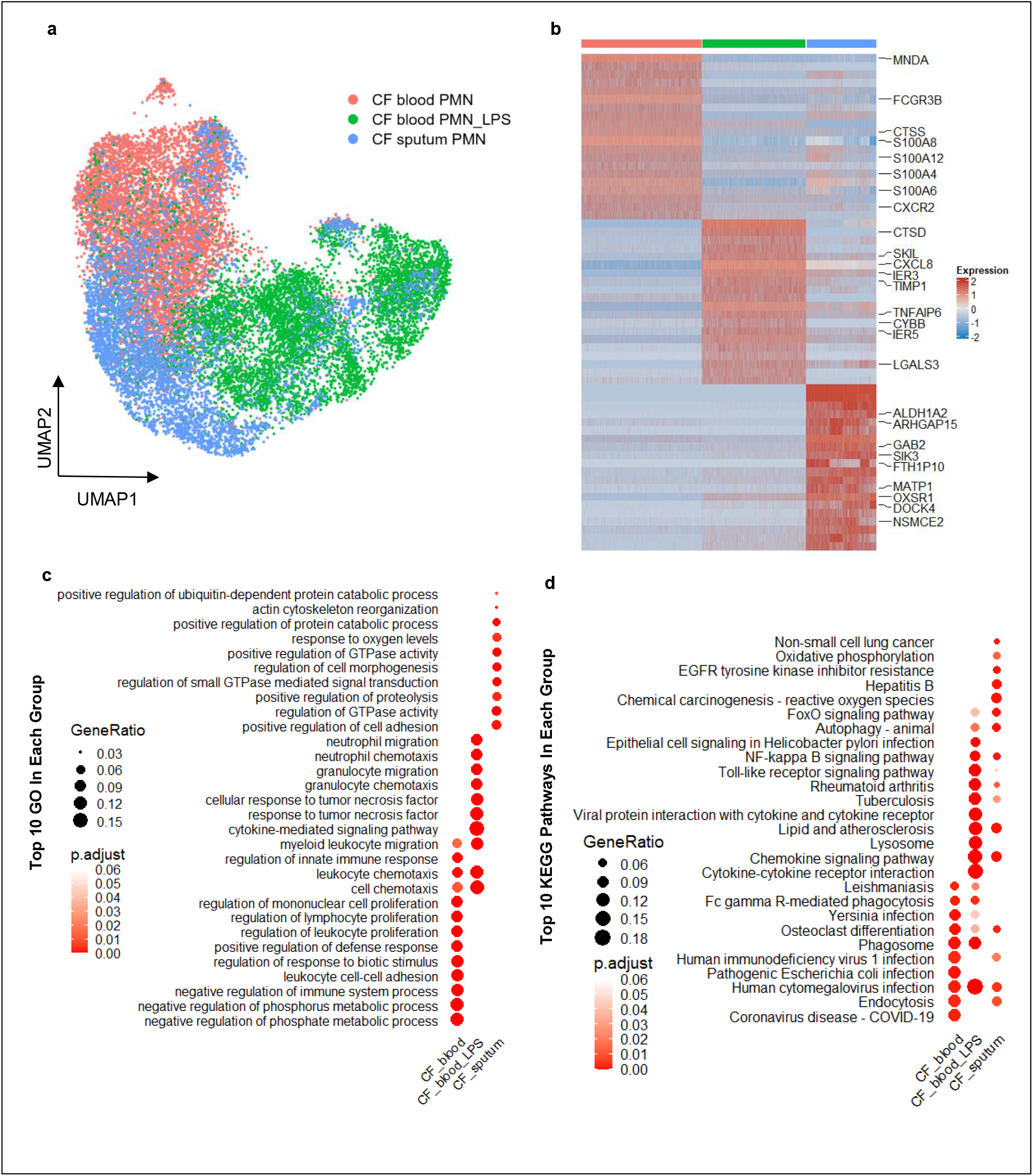
Gene reprogramming in CF sputum neutrophils as compared to CF blood neutrophils. **a**, UMAP of 3 groups of CF PMN: 1) CF blood PMN in their native state, 2) CF blood PMN stimulated with LPS, and 3) CF sputum PMN. **b**, Heat map showing the top 20 DEGs from each group of CF neutrophils. Selected immune-related genes are marked. **c**, GO enrichment comparisons of DEGs among the 3 group of CF neutrophils. **d**, KEGG analysis of DEGs from each group of CF neutrophils.

While a small proportion of CF sputum PMN overlapped with CF blood PMN and CF blood PMN_LPS, CF sputum PMN showed large differences in gene expression (**Fig. 6a & b**). MHC I related genes (*MATP1*), oxidative stress related genes (*OXSR1*), and aldehyde dehydrogenase coding gene *ALDH1A2* were highly expressed in CF sputum PMN (**Fig. 6b**). GO analysis of DEGs revealed that CF sputum PMN shared few GO terms with CF blood neutrophils, suggesting that the CF lung environment imposed significant influences on neutrophil gene programming (**Fig. 6c**). Notably, genes involved in GTPase activity (*ARHGAP15, GAB2*), GTPase activation (*DOCK4*), and proteolysis were on the top GO list from CF sputum PMN. GTPases are known to function as molecular switches to shut down activation(21). Increase in neutrophil proteolysis activity in the lung may cause substantial lung damage. Furthermore, KEGG pathway analysis revealed that CF sputum PMN shared some but not all pathways with CF blood PMN_LPS, including Chemokine signaling pathway, Lipid and atherosclerosis, and NF-κB signaling pathway (**Fig. 6d**).

CF sputum PMN had the lowest differentiation score and highest aging, apoptosis, and necrosis scores (**Fig. 7a**), suggesting that the sputum PMN were in a disordered state leading to incongruent cell activation, survival, and host defense programs. While the cells were less aging relative to circulating PMN, they expressed increased apoptosis- and necrosis-associated genes, when compared with CF blood PMN_LPS samples. CF sputum PMN had significantly lower scores for azurophil, tertiary and secretory granule gene sets, but a significantly higher score for specific granule gene set, as compared to those of CF blood PMN_LPS samples (**Fig. 7b**).

**Fig. 7.**
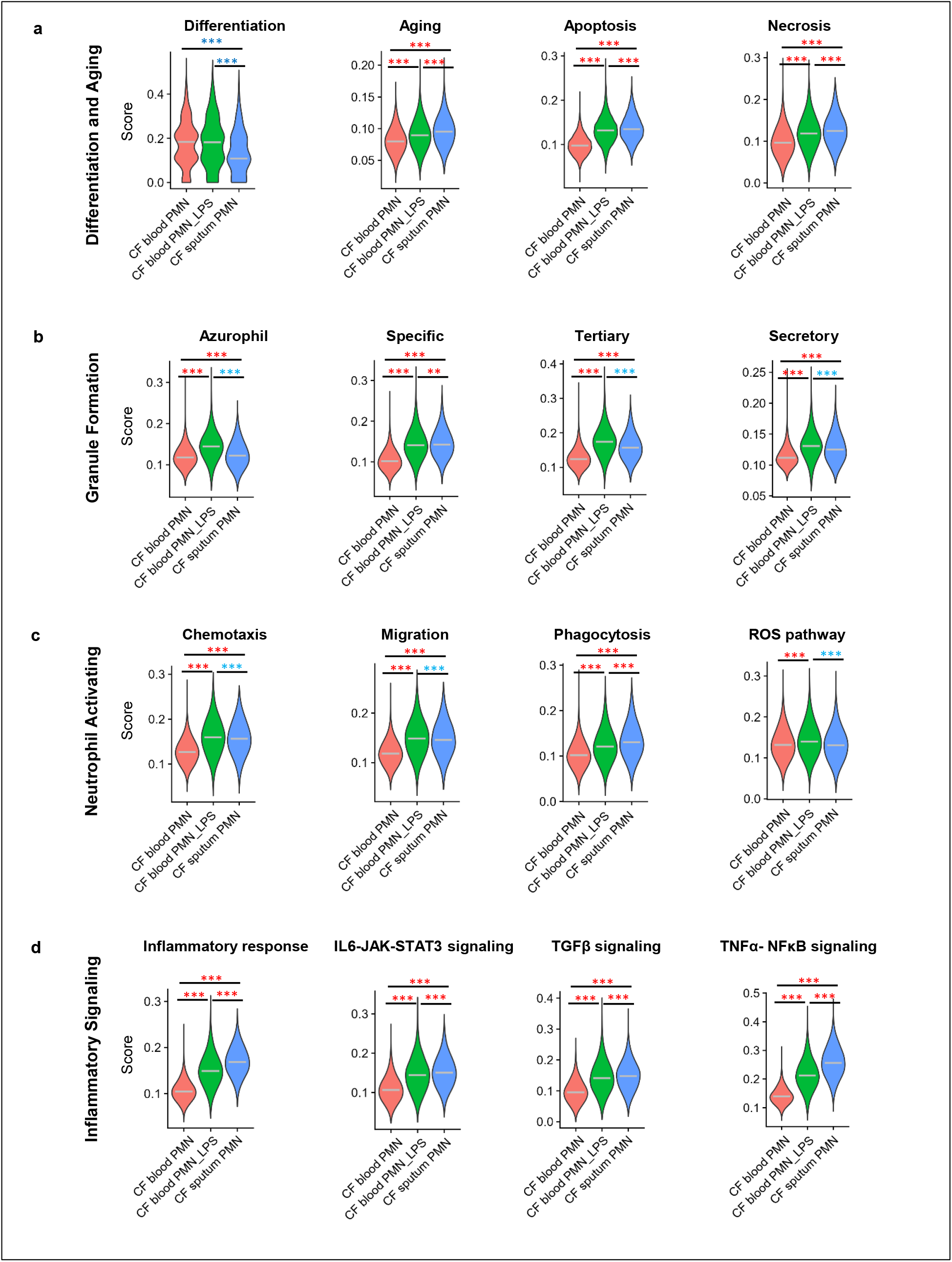
Comparisons of immune-related properties between CF blood neutrophils and CF sputum neutrophils. **a-d**, Comparisons of neutrophil property and function scores among CF blood PMN, CF blood PNM_LPS, and CF sputum PMN. **a**, Differentiation and aging scores; **b**, Granule formation scores; **c**, Neutrophil activation scores; **d**, Inflammation scores. Significance was determined by one-way ANOVA. *p<0.05, **p<0.01, ***p<0.001. Red asterisks indicate that the latter group has significantly higher scores, and blue asterisks significantly lower scores, as compared to the former group in each comparison. Gray lines in each violin plot represent median.

Moreover, the scores for ROS pathway, Migration and Chemotaxis showed a substantial reduction in CF sputum PMN (**Fig. 7c**). Conversely, the score for Phagocytosis was significantly higher in CF sputum PMN (**Fig. 7c**). Furthermore, the gene sets for Inflammatory response, IL6-JAK-STAT3 signaling, TGFβ signaling, and TNFα-NFκB signaling were significantly upregulated in CF sputum PMN (**Fig. 7d**). These comparisons clearly indicated that the immune program in CF sputum neutrophils further deviated from the altered in the CF circulating neutrophils. Taken together, CF sputum neutrophils had a severely disordered transcriptional programming that could delay their maturation, exacerbate immune dysfunction and accelerate necrotic cell death. These data provide one molecular explanation to how CF lungs are inflicted by persistent neutrophilic inflammation and purulent airway obstructions.

## Discussion

Persistent neutrophilic inflammation is a major pathogenic factor in CF that damages organ structure and function. However, the mechanism underlying the pathogenesis has not been clearly defined. The current report provides comprehensive data demonstrating that CF neutrophils have intrinsic anomalies in immune programming. Notably, peripheral blood neutrophils from CF patients exhibited pro-inflammatory priming before recruitment to any target organs. Such dysregulated and premature activation led to an exaggerated but dysregulated immune response at the transcriptional level. Of note, our CF subjects were all under the Elexacaftor/Tezacaftor/Ivacaftor modulator therapy. Such observed abnormalities suggest that the current CFTR modulators may not be effective in rectifying CF neutrophil innate immune defects.

An early microarray analysis revealed that peripheral blood neutrophils from CF patients upregulate their pro-inflammatory genes, including *G-CSF, CXCL10, CCL17, IKKɛ*, and *IL-10Ra* (22), which is consistent with our finding that CF circulating neutrophils are pre-primed. Schupp and colleagues performed scRNA-seq of neutrophils isolated from the sputum of CF patients, and found a prevalence of immature pro-inflammatory neutrophils(20). Direct comparison of their CF sputum neutrophil data with our CF blood neutrophil data has revealed significant transcriptional re-programming, which amplified the innate abnormality of CF circulating neutrophils. These data have confirmed a recently published bulk-RNA-seq study that demonstrated that CF neutrophils, after migrating through an air-liquid interface culture of H441-cell-derived airway epithelia, undergo extensive transcriptional transformations related to immune response genes (23).

Numerous previous reports have documented CF PMN dysfunction phenotypically, including suboptimal activation (24), cleavage of CXCR1 (25), hyper-sensitivity to LPS stimulation (26), irregular or deficient production of reactive oxygen species (27, 28), alteration in inflammatory signaling (29), hyper-production of IL-8 (30, 31), abnormal extracellular trap formation (32), hyper-oxidation of glutathione (33), dysregulation of protease secretion and degranulation, delayed apoptosis (34), and abnormal granule release (35). Moreover, CF blood neutrophils express higher levels of CD64, a marker of neutrophil activation, and lower levels of Toll-like receptor-2 (TLR2) compared with healthy blood neutrophils. Further, TLR2, TLR4 and IL-8 expression in CF airway neutrophils was higher than in CF blood neutrophils (36). As cell phenotype is at least in part determined by gene activities, these phenotypic abnormalities reflect gene programming anomalies in CF neutrophils. Our current scRNA-seq data will be valuable in understanding the molecular basis of these functional defects.

In this report, three tiers of comparisons of neutrophil transcriptomes were performed: 1) inter-genotype comparison (HC vs CF); 2) inter-state comparison (basal state vs activating state vs aging state) of each genotype; and 3) inter-compartment comparison (blood vs sputum). The inter-genotype comparison allowed us to discern the differences in gene programming between genotypes. We found that HC and CF neutrophils shared many core expressing genes (**Fig. 1d**), but CF neutrophils in the blood were innately primed (**Fig. 1f & 2a**). Such a premature activation is independent of external inductions by pro-inflammatory cytokines or endotoxin. More importantly, despite the early start in activation CF neutrophils failed to organize an effective immune response to LPS at the transcriptional level.

The inter-state comparison uncovered how CF and HC neutrophils responded to LPS challenge differently. We found that CF neutrophils differed from HC neutrophils in their fate decision and aging/death processes. Instead of natural aging and apoptosis in HC neutrophils, CF neutrophils manifested a senile activation. This abnormal aging process might lead to a prolonged state of neutrophilic inflammation. Moreover, CF neutrophils had a significantly higher necrosis score in either activating or aging state (**Fig. 4g**), and a significantly higher combined necrosis score of both states (**Supplementary Fig. 7**). Necrosis is a cell autolysis that does not follow the apoptotic signaling pathway, from which uncontrolled release of products of cell death into the extracellular space provokes strong inflammatory response (37). The high necrosis score of CF neutrophils upon LPS challenge may underlie a mechanism for purulent inflammation observed in CF lungs.

The inter-compartment comparison revealed that CF sputum neutrophils had significantly less enrichment of gene transcripts related to azurophil and tertiary granule formation and ROS production, when compared with LPS-stimulated CF circulating neutrophils. Neutrophil granules play an essential role in neutrophil innate immune function. Four types of granules are produced during neutrophil development and maturation (11). Primary granules are formed at the earliest stage of neutrophil differentiation in the bone marrow, and contain potent proteases, such as neutrophil elastase (NE), cathepsin G, the chlorinating enzyme myeloperoxidase (MPO) and antimicrobial agents such as defensins (38). Although LPS stimulation upregulates the different granule genes in healthy neutrophils (11), which was also proved in our data (**Supplementary Fig. 7**), CF blood neutrophils under LPS challenge had even higher expression of these genes (**Fig. 4h**). Our analyses have also shown that CF sputum neutrophils had significantly higher granule scores even than the LPS-challenged CF blood neutrophils (**Fig. 7b**), suggesting that CF lungs may have more abundant or stronger agonists to stimulate the lung-recruited neutrophils.

Primary granules generally fuse with the phagosome for bacterial killing to avoid tissue damage (38, 39), but uncontrolled degranulation of CF neutrophils does not serve anti-microbial function, instead produces bystander tissue damage (23). Large amounts of NE and MPO have been found in the airway at a very early life stage of CF patients (40). NE burden in the airway accelerated the cleavage of CXCR1, resulting in poor bacterial killing but persistent inflammation. NE activity is negatively correlated with lung function in CF patients (41). Thus, an unbalanced production of granules would interfere with optimal phagosomal function and effective lung defense.

Human neutrophils pre-synthesize CFTR that is pre-stored in the secretory vesicle membrane (28). When integrated into the phagosomal membrane, CFTR serves as a major chloride channel to transport chloride to the organelle (42). CFTR is crucial to regulating phagosomal functions, including charge balance, pH maintenance and oxidant production (43-45). Our previous work and others’ have demonstrated that CFTR loss of function in CF neutrophils results in a deficit in chloride supply to the phagosomes, leading to deficiency in microbial killing and resolution of inflammation (42, 46, 47). In addition to the phagosomal chloride supply and oxidant production defect, CF neutrophils have aberrant gene programming, which may further undermine neutrophil innate immune function.

In summary, our study has provided critical data demonstrating that CF neutrophils are intrinsically pro-inflammatory. Such an innate anomaly is determined by internal gene programming, which renders the cells hyper-responsive to stimulation, and less effective in host defense function. This finding helps explain at the gene level why CF lungs have unremitting neutrophilic inflammation that causes long-term tissue damage and accelerated lung functional decline in CF.

This study has several limitations: First, the sample sizes of CF and HC cohorts were relatively small. As we only targeted the single neutrophil population, deep sequencing of enough high-quality cells was readily achieved. However, only 3 CF and 3 HC subjects were recruited to this study. Second, this is a single center investigation. Although published datasets of CF and HC subjects from other centers were mined and compared with ours, our own sequencing data were from the participants from a single CF care center. Third, all the recruited CF patients were homozygous in F508del CFTR gene mutation, and had been receiving the ETI modulator therapy. Patients with other CFTR mutations and receiving no modulator treatments were not covered in this study.

## MATERIALS AND METHODS

### Human study ethical statement

This study involved human blood samples. The experimental design was approved by the Institutional Review Boards of Tulane University Health Sciences Center and Louisiana State University Health Sciences Center, Louisiana State, USA. Informed consent from each participant was obtained before any sample collection.

### Human neutrophil isolation and processing

Peripheral blood (10 cc) from 3 CF patient and 3 healthy donors (**Supplementary Table 1**) were drawn into a BD Vacutainer® Blood Collection Tube (EDTA K2). The selected patients were homozygous for the F508del variant or ΔF508) with stable mild to moderate lung disease. Lung disease stability was clinically defined as demonstrating no recent change in lung function and without clinical signs of acute bronchiectasis exacerbation. All 3 CF participants were being treated with the CFTR protein modulators: elexacaftor/texacaftor/ivacaftor (ETI). After Percoll-gradient centrifugation (65%/75%), the neutrophil layer of each sample was collected and resuspended in pre-cooled RPMI-1640 (Gibco) medium without any supplements. The obtained cells were divided into two aliquots: one immediately processed for scRNA-seq, and the other cultured in complete RPMI-1640 media supplemented with 2 mM GlutaMax (Gibco), 10% heat-inactivated bovine growth serum (Hyclone), 100 U/ml penicillin (Gibco), 100 μg/ml streptomycin (Gibco) and 0.25 μg/ml amphotericin B (Sigma), and stimulated with *P. aeruginosa* LPS (Sigma; 40 µg/mL) for 16 hours, followed by scRNA-seq.

### Cell hashing, library construction and sequencing

In order to minimize sample to sample variations, hashtag oligonucleotides (HTO) against human β2-microglobulin (β2M) were used to tag different groups of cells. Three samples from blood donors in each group were tagged with following hashtag oligonucleotides: TotalSeq™-B0251 anti-human Hashtag 1 Antibody (BioLegend Cat. No. 394601), TotalSeq™-B0252 anti-human Hashtag 2 Antibody (BioLegend Cat. No. 394603); TotalSeq™-B0253 anti-human Hashtag 3 Antibody (BioLegend Cat. No. 394605). Cells were loaded onto the 10X Chromium controller, and cell viability was evaluated by Cellometer (Nexcelom Bioscience) with AO/PI staining. Five thousand neutrophils from each donor were targeted. The libraries were prepared using Chromium Single Cell 3ʹ v3.1 protocol (10x Genomics). Cellular mRNA and antibody-derived oligos were reverse-transcribed and indexed with a shared cellular barcode. The mRNA derived cDNA and antibody-derived tagged DNA were constructed to libraries according to the Chromium Single Cell 3’ v3.1 protocol (10x Genomics). The mRNA derived cDNA library and antibody-derived tagged library were pooled and sequenced at 4:1 ratio with paired-end reads on an Illumina NextSeq 2000 (Illumina).

### HL-60 cell culture and neutrophil differentiation

A homozygous F508del-CF HL-60 cell line (CF_HL-60)(48) and the parental wild-type HL-60 cell line (WT_HL-60) were cultured in RPMI-1640 (Gibco) supplemented with 2 mM GlutaMax (Gibco), 10% heat-inactivated bovine growth serum (Hyclone), 100 U/ml penicillin (Gibco), 100 μg/ml streptomycin (Gibco) and 0.25 μg/ml amphotericin B (Sigma). To differentiate the cells into neutrophils, 1.25% of DMSO (Sigma-Aldrich) was added to the medium for 5 days with medium replenishment at Day 3. The differentiated neutrophils were submitted for scRNA-seq using the same procedure as for the human blood neutrophils described above.

### ScRNA-seq data processing

Primary analyses were performed using 10x Cellranger V.6.1.2, including alignment of the sequencing reads to the GRCh38-2020-A human transcriptome, and quantification of transcript expressions in each cell. The downstream quality control and data analyses were achieved using R version 4.1.3 and the package Seurat version 4.2.1(49) except where otherwise noted.

#### Seurat data QC and demultiplex

The output folder from Cell Ranger 6.1.2, containing barcodes, features, and matrix, was loaded into R by the Read10X() function from Seurat. For quality control, cells that have mitochondrial gene counts lower than 5% with unique feature counts more than 150 and less than 2500 (subset = percent.mt < 5 & nFeature_RNA>150 & nFeature_RNA<2500)), were accepted for downstream analysis. All the qualified cells were demultiplexed using the HTODemux() function, according to their HTO barcode. Doublet and negative cells were ruled out.

#### Dimension reduction and unsupervised clustering

After removing unwanted (low quality) cells, the dataset was normalized by the NormalizeDate() function, and scaled by the SCTransform() function with *method =* “*glmGamPoi*”, *vars*.*to*.*regress =* “*percent*.*mt*”. The first 30 principal components (PCs) were used to compute a Uniform Manifold Approximation and Projection (UMAP) embedding of the cells with the functions: FindNeighbors(), RunUAMP(), and FindClusters(). The states of neutrophils were identified by Monocle2 2.24.1(50) using the reduceDimension() function. To integrate datasets and eliminate the batch-batch difference between each dataset, R package Harmony 0.1.0 was used for batch correction (51).

#### Identification of differentially expressed genes (DEGs) and analysis of gene ontology (GO)

Differentially expressed genes of each cluster or group were identified by function FindMarkers() or FindAllMarkers() with adjusted P value < 0.05 and log2FC > 0.5. Bonferroni correction was performed to adjust P value. The lists of DEGs were also used for GO analysis and KEGG pathway analysis by using the R package ClusterProfiler 4.2.2 (52).

#### RNA velocity analysis

RNA velocity analysis was performed using velocyto.py 0.17.16 (15) command-line tools to convert bam files to loom files. Then, the downstream analysis was performed by scVelo 0.2.5 pipeline (53) to estimate RNA velocity at dynamic states (scv.tl.recover_dynamics). We used Seurat cells embedding to show the RNA velocity, root cells, and end point of the underlying cellular processes.

#### Scoring of biological process and pathway activity

Individual cells were scored for their expression of gene signatures representing certain biological functions and pathway activities. Each functional signature gene set was derived from the Molecular Signatures Database (MSigDB), including hallmark gene sets (54) and the Gene Ontology gene sets. Neutrophil function related scores (GO: 0042119; 0030593; 0019221; 1990266; 0002764; 0002224; 0002221; 0032496) were calculated by the AddModuleScore () function with DEGs enriched in corresponding GO terms. All other signature functional scores reflecting certain gene set activity were calculated by R package: AUCell 1.18.1 (55) with the AUCell_buildRankings() function to compute gene expression rankings in each cell and the AUCell_calcAUC() function to calculate area-under-the-curve (AUC) values of each gene set in a cell. AUC values represent the fraction of genes within the top-ranking genes for each cell that are defined as part of the pathway gene set (56).

### Cytokine and endotoxin measurements

R&D Systems DuoSet ELISAs were used to assess TNF-α, IL-8, IL-6, and IL-1β levels in healthy control plasma samples and CF patient plasma samples according to the manufacturer protocol with freshly made buffers and diluents. In brief, plates were coated overnight with capture antibody reconstituted in PBS (Gibco). Plates were washed and blocked for 2 hours. Standards and samples were then prepared and diluted in proper diluents as recommended by the manufacturer and incubated at room temperature for 2 hours. Plates were washed with 0.05% Tween-20 (Thermo Fisher) in PBS. Detection antibody reconstituted in appropriate reagent diluent as per manufacturer protocol was added to samples and standards and incubated at room temperature for 2 hours. Plates were washed again. Working dilutions of Streptavidin-HRP (R&D Systems) were added to each well and incubated in the dark for 20 min. Plates were washed and substrate solution (Thermo Scientific Ref: 34028) was added to each well and plates were incubated in the dark for 20 min. Stop solution (2N H_2_SO_4_) was added to each well and plates were measured at 540 nm and 450 nm. Wavelength correction was performed by subtracting readings at 540 nm from readings at 450 nm.

For endotoxin measurement, the Pierce Chromogenic Endotoxin Quant Kit (Thermo Scientific Cat: A39552) was used to determine endotoxin levels in healthy human and CF patient plasma samples according to the manufacturer instructions. All reagents were pre-warmed and kept at 37°C using a plate warmer. In brief, endotoxin standards were made by reconstituting lyophilized endotoxin with endotoxin free water. Standards with low concentrations (0.01-0.1 EU/mL) were made according to the recommended protocol. Plasma samples were diluted 50 fold in endotoxin free water per protocol suggestion. Standards and samples were added to each well prior to addition of reconstituted Amebocyte Lysate Reagent. Samples were incubated at 37°C for 20 min (time directed as T1 for Low standard time). Reconstituted chromogenic substrate solution was added to each well and incubated at 37°C for 6 min. Stop solution (25% acetic acid) was added to each well and the plate was read at 405 nm. To validate the measurement, randomly selected plasma samples were spiked with a known amount of LPS, and measured in parallel.

### Statistical analyses

A two-tailed, unpaired Student’s t-test was used for 2 group comparisons. For one factor, more than 2 group comparisons, one-way ANOVA was performed. For 2 factors comparisons, two-way ANOVA was performed. The values shown in each figure are represented as mean ± SD. P<0.05 was considered statistically significant (*p<0.05, **p<0.01. ***p<0.001). All statistical analyses and graphics were made using GraphPad Prism 9, SigmaStat (SyStat Software, San Jose, CA), or R 4.1.3 (package: ggplot2 3.3.6).

## Data Availability

All scRNA-seq data have been deposited in the Gene Expression Omnibus (GEO) database. The human blood neutrophil data (CF patients and healthy controls) are under the accession code (GSE220658), and the HL-60-derived neutrophil data are under the accession code (GSE220913).

## Data availability

All scRNA-seq data have been deposited in the Gene Expression Omnibus (GEO) database. The human blood neutrophil data (CF patients and healthy controls) are under the accession code GSE220658, and the HL-60-derived neutrophil data are under the accession code GSE220913. This study has also mined other investigators’ published datasets: 1) the healthy control blood neutrophil data from the Franco Lab (14) are under the accession code GSE188288, and the CF sputum neutrophil data from Britto Lab (20) are under the accession code GSE145360. All the used subjects from the two published datasets were selected to match the criteria of our human subjects.

## Code availability

No custom software was used in this study. Methods described all the used analytical tools. R code is available from the corresponding author upon request.

## Acknowledgments

The authors would like to thank Dr. William M. Nauseef at University of Iowa for his advice on this project. This work was supported by grants to GW from The National Institutes of Health (HL150370) and The Cystic Fibrosis Foundation (WANG18I0) and by the Gilead Research Scholar Program in Cystic Fibrosis to CMB. Portions of this research were conducted with high performance computational resources provided by the Louisiana Optical Network Infrastructure (http://www.loni.org).

## Author contributions

YH, DW, KS, CS and SC performed the experiments; CMB did clinical evaluation, subject selection and consent, and blood sample collection; KS and JKK performed the single-cell RNA sequencing and primary data analyses. YH, JL and CJB carried out the secondary data analyses.

