## Supplementary Information for "ABERRANT IMMUNE PROGRAMMING IN NEUTROPHILS IN CYSTIC FIBROSIS"

**Supplementary Table 1: Demographics of CF participants and HC donors**

| Subjects | CF | | HC | |
|---|---|---|---|---|
| Dataset | Wang Lab GSE220658 (n=3) | Britto Lab GSE145360 (n=6) | Wang Lab GSE220658 (n=3) | Franco Lab GSE118288 (n=6) |
| Age, yr | 20-37 | 24-37 | 27-58 | 21-33 |
| Sex | 1 female; 2 male | 3 female; 3 male | 2 female; 1 male | 2 female; 4 male |
| Mutation Backgroud | F508del/F508del | F508del/F508del | N/A | N/A |
| $FEV_1$, L | 1.81-3.7 | 0.68–2.85 | N/A | N/A |
| $FEV_1$, % | 59-98 | 19-65 | N/A | N/A |
| Culture History | | | | |
| *P. aeruginosa* | 2 | 6 | N/A | N/A |
| *S. aureus* | 2 | N/A | N/A | N/A |
| CFTR modulators | ETI | Yes | N/A | N/A |

### Supplementary Figure 1

**a**

| Barcode | Sample name |
|---|---|
| hPMN-B0251 | HC_1 |
| hPMN-B0252 | HC_2 |
| hPMN-B0253 | HC_3 |

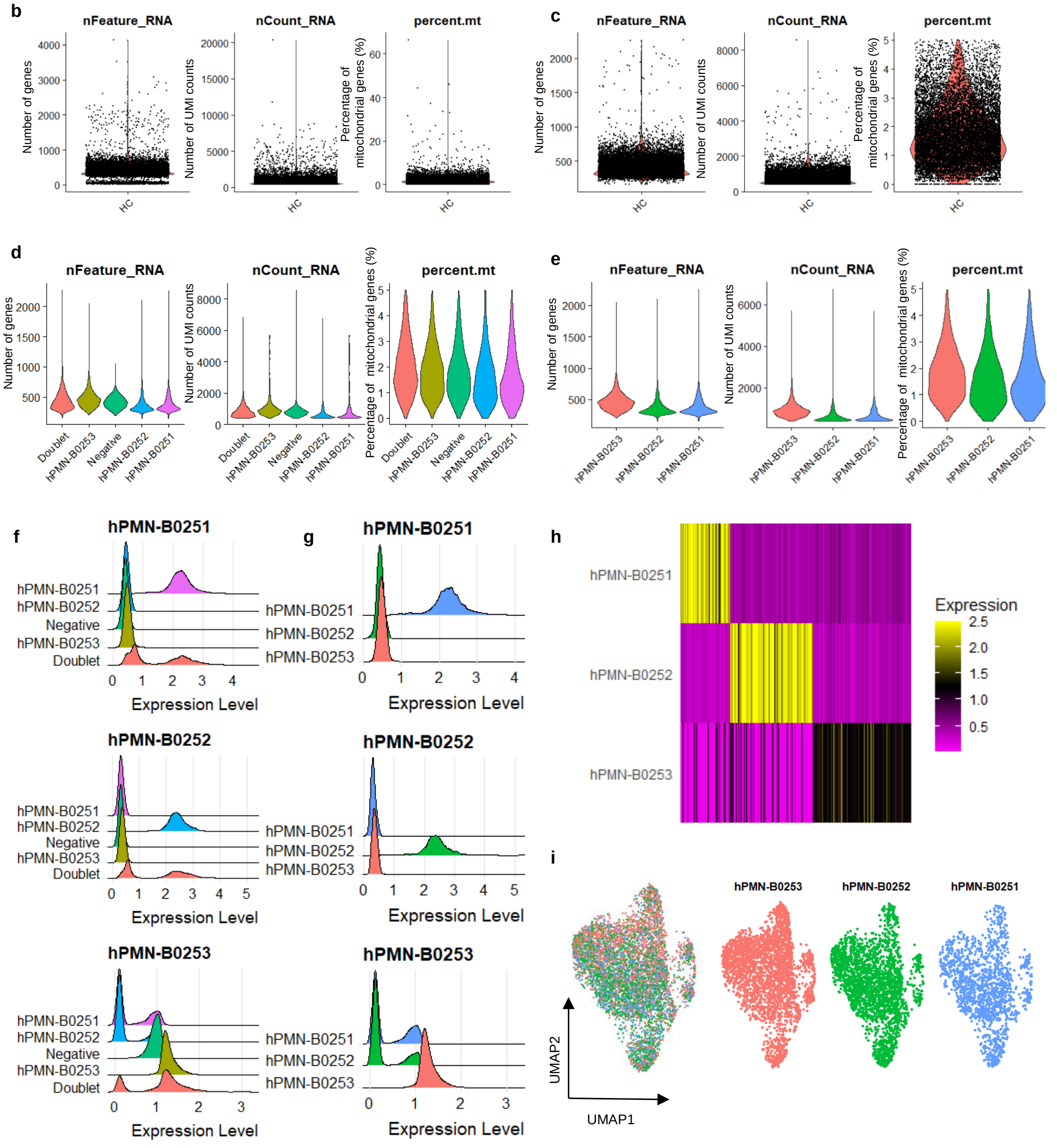

**Supplementary Fig. 1. ScRNAseq data quality control for HC samples. a**, Sample information and hashtag oligonucleotide (HTO) barcodes. **b**, Violin plots of gene counts (nFerature_RNA, left panel), UMI count (nCount_RNA, middle panel), and percentage of mitochondrial gene counts (percent.mt, right panel) of the data set before quality control. **c**, Violin plots of gene counts (nFerature_RNA, left panel), UMI count (nCount_RNA, middle panel), and percentage of mitochondrial gene counts (percent.mt, right panel) of the data set after quality control filtering. **d**, Violin plots of gene counts (nFerature_RNA, left panel), UMI count (nCount_RNA, middle panel), and percentage of mitochondrial gene counts (percent.mt, right panel) from each sample before demultiplexing. **e**, Violin plots of gene counts (nFerature_RNA, left panel), UMI count (nCount_RNA, middle panel), and percentage of mitochondrial gene counts (percent.mt, right panel) from the singlets of each demultiplexed sample. **f**, Ridge plots of HTO samples enriched before demultiplexing. **g**, Ridge plots of singlets from HTO sample enriched after demultiplexing. **h**, Heamap showing singlets from HTO sample enriched after demultiplexing. **i**, UMAP of HC samples after quality control and demultiplexing, colored by 3 samples together or split by HTO.

### Supplementary Figure 2

**a**

| Barcode | Sample name |
|---|---|
| hPMN-B0251 | HC_LPS_1 |
| hPMN-B0252 | HC_LPS_2 |
| hPMN-B0253 | HC_LPS_3 |

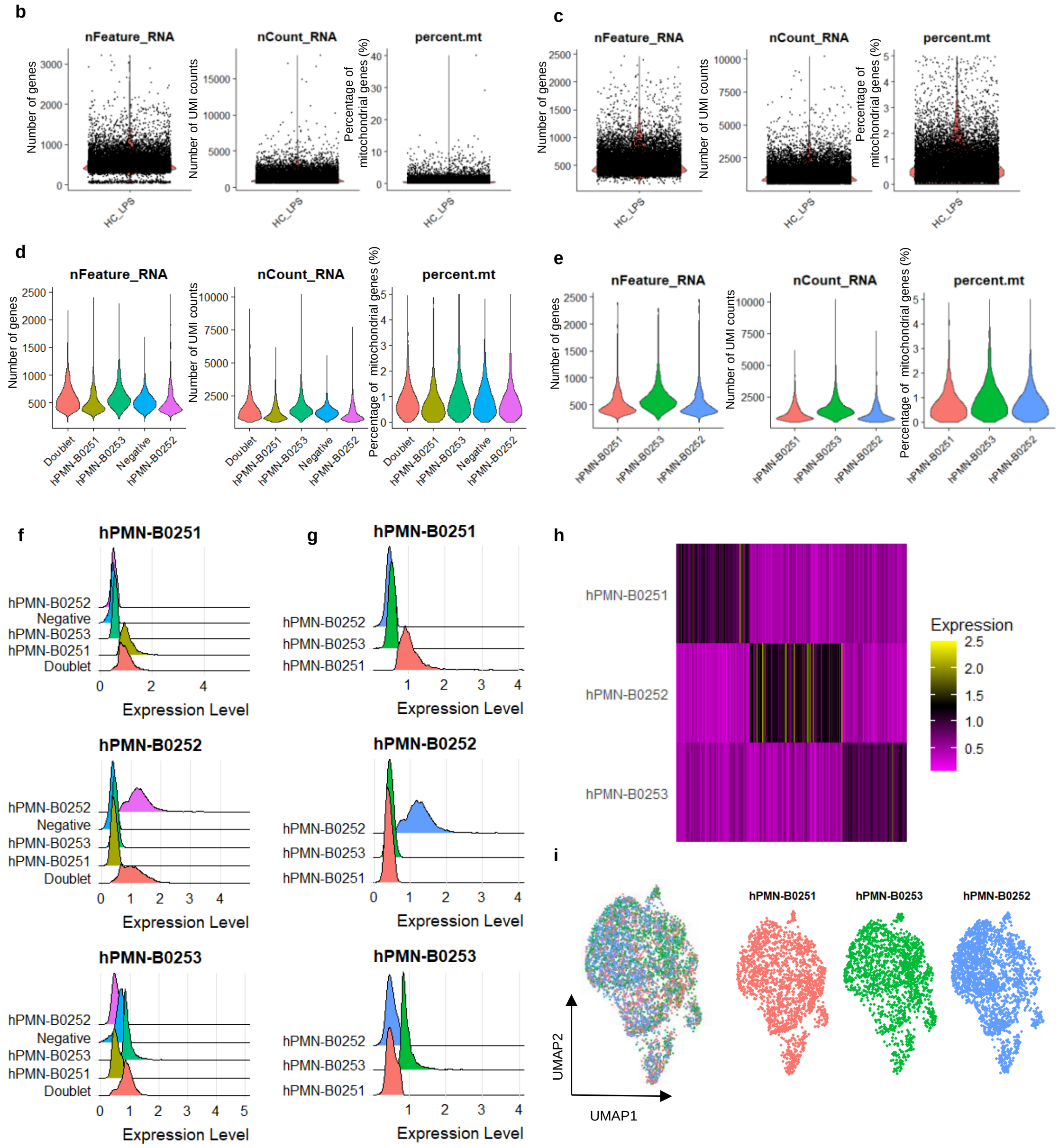

**Supplementary Fig. 2. ScRNAseq data quality control for HC_LPS samples. a**, Sample information and hashtag oligonucleotide (HTO) barcodes. **b**, Violin plots of gene counts (nFerature_RNA, left panel), UMI count (nCount_RNA, middle panel), and percentage of mitochondrial gene counts (percent.mt, right panel) of the data set before quality control. **c**, Violin plots of gene counts (nFerature_RNA, left panel), UMI count (nCount_RNA, middle panel), and percentage of mitochondrial gene counts (percent.mt, right panel) of the data set after quality control filtering. **d**, Violin plots of gene counts (nFerature_RNA, left panel), UMI count (nCount_RNA, middle panel), and percentage of mitochondrial gene counts (percent.mt, right panel) from each sample before demultiplexing. **e**, Violin plots of gene counts (nFerature_RNA, left panel), UMI count (nCount_RNA, middle panel), and percentage of mitochondrial gene counts (percent.mt, right panel) from the singlets of each demultiplexed sample. **f**, Ridge plots of HTO samples enriched before demultiplexing. **g**, Ridge plots of singlets from HTO sample enriched after demultiplexing. **h**, Heamap showing singlets from HTO sample enriched after demultiplexing. **i**, UMAP of HC samples after quality control and demultiplexing, colored by 3 samples together or split by HTO.

### Supplementary Figure 3

**a**

| Barcode | Sample name |
|---|---|
| hPMN-B0251 | CF_1 |
| hPMN-B0252 | CF_2 |
| hPMN-B0253 | CF_3 |

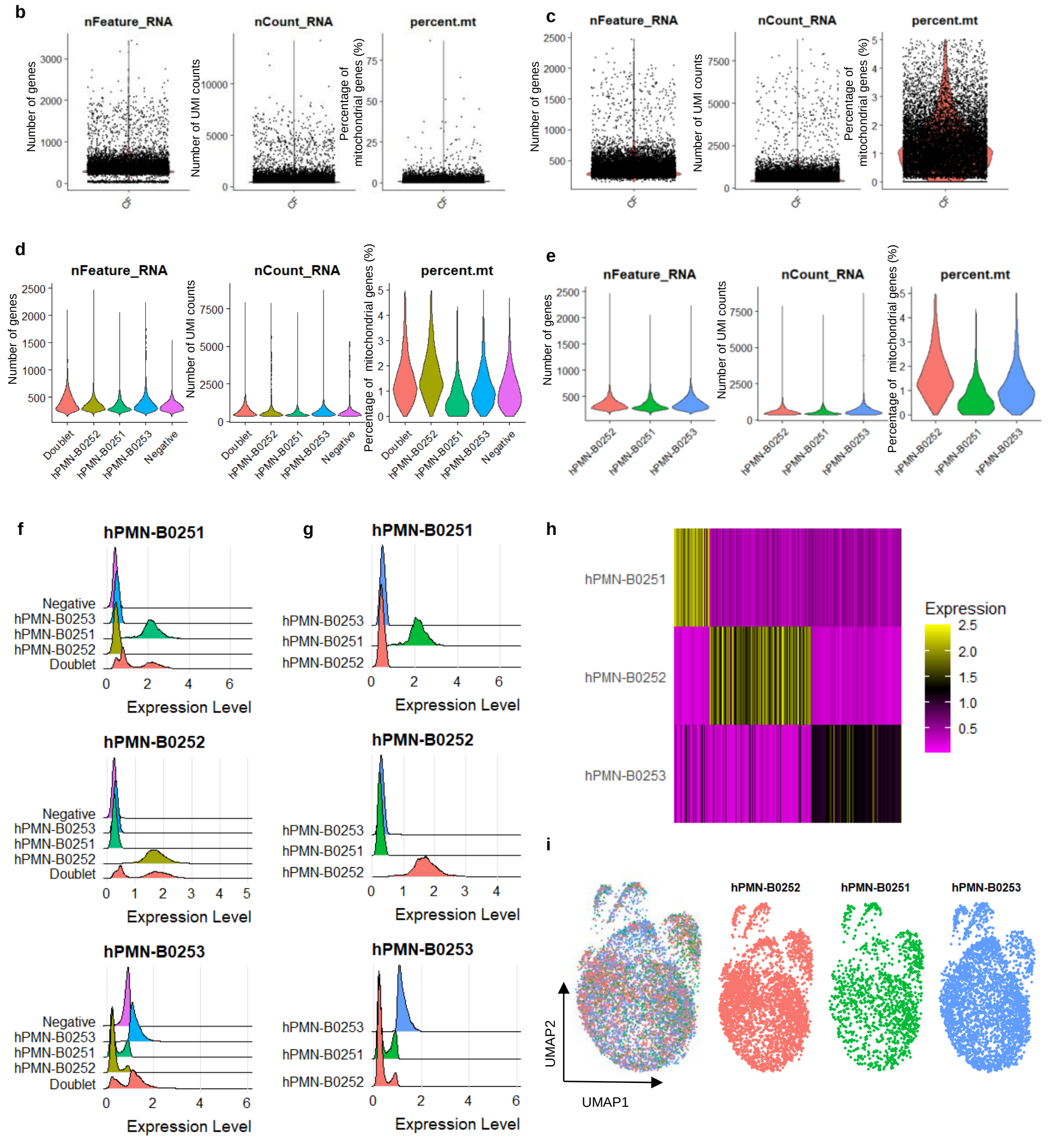

**Supplementary Fig. 3. ScRNAseq data quality control for CF samples. a**, Sample information and hashtag oligonucleotide (HTO) barcodes. **b**, Violin plots of gene counts (nFerature_RNA, left panel), UMI count (nCount_RNA, middle panel), and percentage of mitochondrial gene counts (percent.mt, right panel) of the data set before quality control. **c**, Violin plots of gene counts (nFerature_RNA, left panel), UMI count (nCount_RNA, middle panel), and percentage of mitochondrial gene counts (percent.mt, right panel) of the data set after quality control filtering. **d**, Violin plots of gene counts (nFerature_RNA, left panel), UMI count (nCount_RNA, middle panel), and percentage of mitochondrial gene counts (percent.mt, right panel) from each sample before demultiplexing. **e**, Violin plots of gene counts (nFerature_RNA, left panel), UMI count (nCount_RNA, middle panel), and percentage of mitochondrial gene counts (percent.mt, right panel) from the singlets of each demultiplexed sample. **f**, Ridge plots of HTO samples enriched before demultiplexing. **g**, Ridge plots of singlets from HTO sample enriched after demultiplexing. **h**, Heamap showing singlets from HTO sample enriched after demultiplexing. **i**, UMAP of HC samples after quality control and demultiplexing, colored by 3 samples together or split by HTO.

### Supplementary Figure 4

**a**

| Barcode | Sample name |
|---|---|
| hPMN-B0251 | CF_LPS_1 |
| hPMN-B0252 | CF_LPS_2 |
| hPMN-B0253 | CF_LPS_3 |

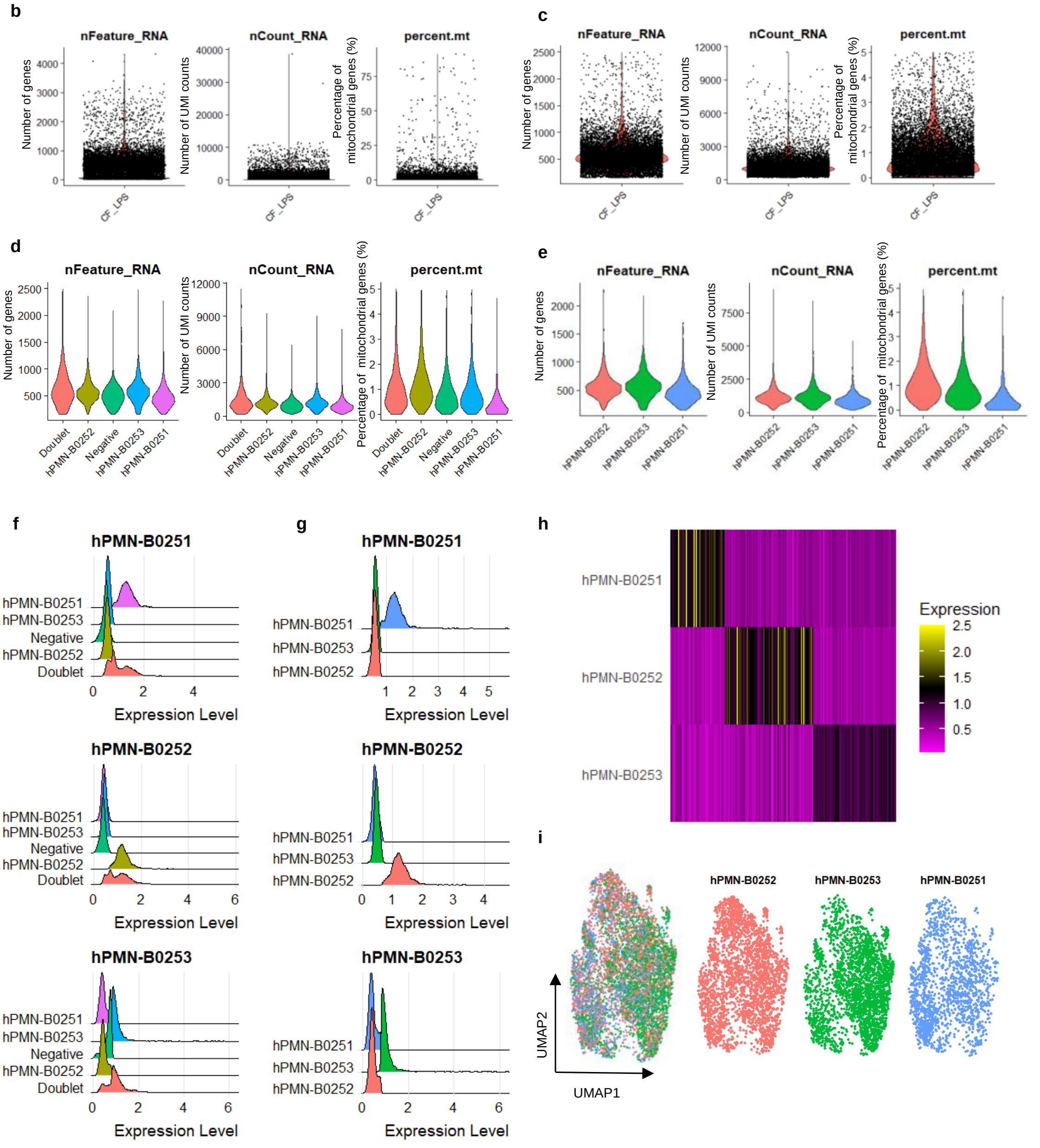

**Supplementary Fig. 4. ScRNAseq data quality control for CF_LPS samples. a**, Sample information and hashtag oligonucleotide (HTO) barcodes. **b**, Violin plots of gene counts (nFerature_RNA, left panel), UMI count (nCount_RNA, middle panel), and percentage of mitochondrial gene counts (percent.mt, right panel) of the data set before quality control. **c**, Violin plots of gene counts (nFerature_RNA, left panel), UMI count (nCount_RNA, middle panel), and percentage of mitochondrial gene counts (percent.mt, right panel) of the data set after quality control filtering. **d**, Violin plots of gene counts (nFerature_RNA, left panel), UMI count (nCount_RNA, middle panel), and percentage of mitochondrial gene counts (percent.mt, right panel) from each sample before demultiplexing. **e**, Violin plots of gene counts (nFerature_RNA, left panel), UMI count (nCount_RNA, middle panel), and percentage of mitochondrial gene counts (percent.mt, right panel) from the singlets of each demultiplexed sample. **f**, Ridge plots of HTO samples enriched before demultiplexing. **g**, Ridge plots of singlets from HTO sample enriched after demultiplexing. **h**, Heamap showing singlets from HTO sample enriched after demultiplexing. **i**, UMAP of HC samples after quality control and demultiplexing, colored by 3 samples together or split by HTO.

**Supplementary Figure 5**

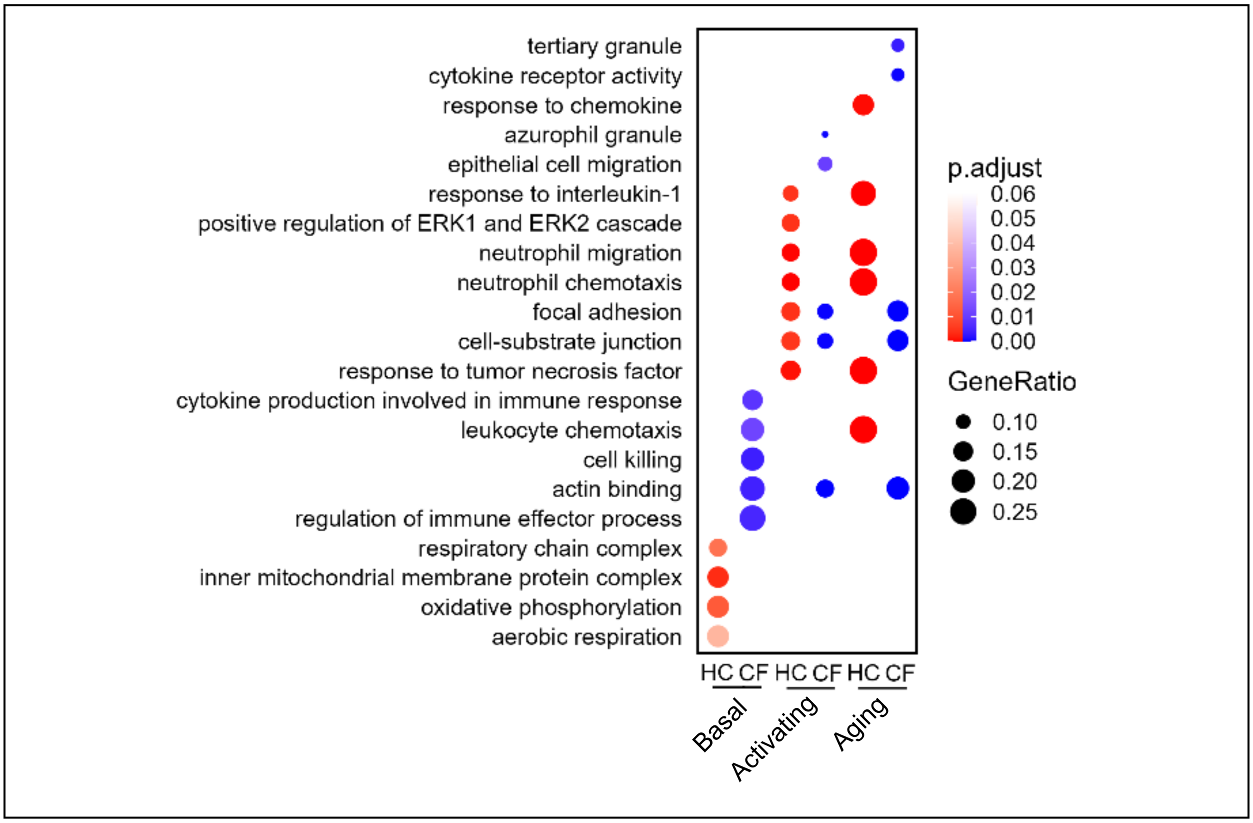


**Supplementary Fig. 5. Gene Ontology enrichment of DEGs of genotype comparison in each state.** The dot plot shows two-group comparison of GO between healthy control (HC) and cystic fibrosis (CF) groups in their basal, activating, and aging states. Red dots belong to HC group, and blue dots belong to CF groups. The size of dot represents the gene ratio, and the gradient change of colors represents adjusted p value.

**Supplementary Figure 6**

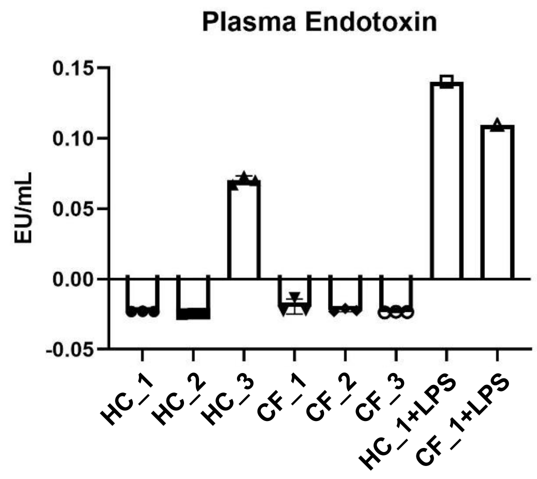


**Supplementary Fig. 6. Plasma endotoxin levels of HC and CF samples.** Plasma endotoxin levels in HC and CF samples were assessed using the Thermo Scientific endotoxin kit. Samples were run in triplicate and measured against the kit-provided Low EU standard. To confirm veracity of the method, HC1 and CF1 plasma samples were spiked with 0.10 EU/mL of exogenous LPS (HC_1+LPS, CF_1+LPS), and measured in parallel as a control.

#### Supplementary Figure 7

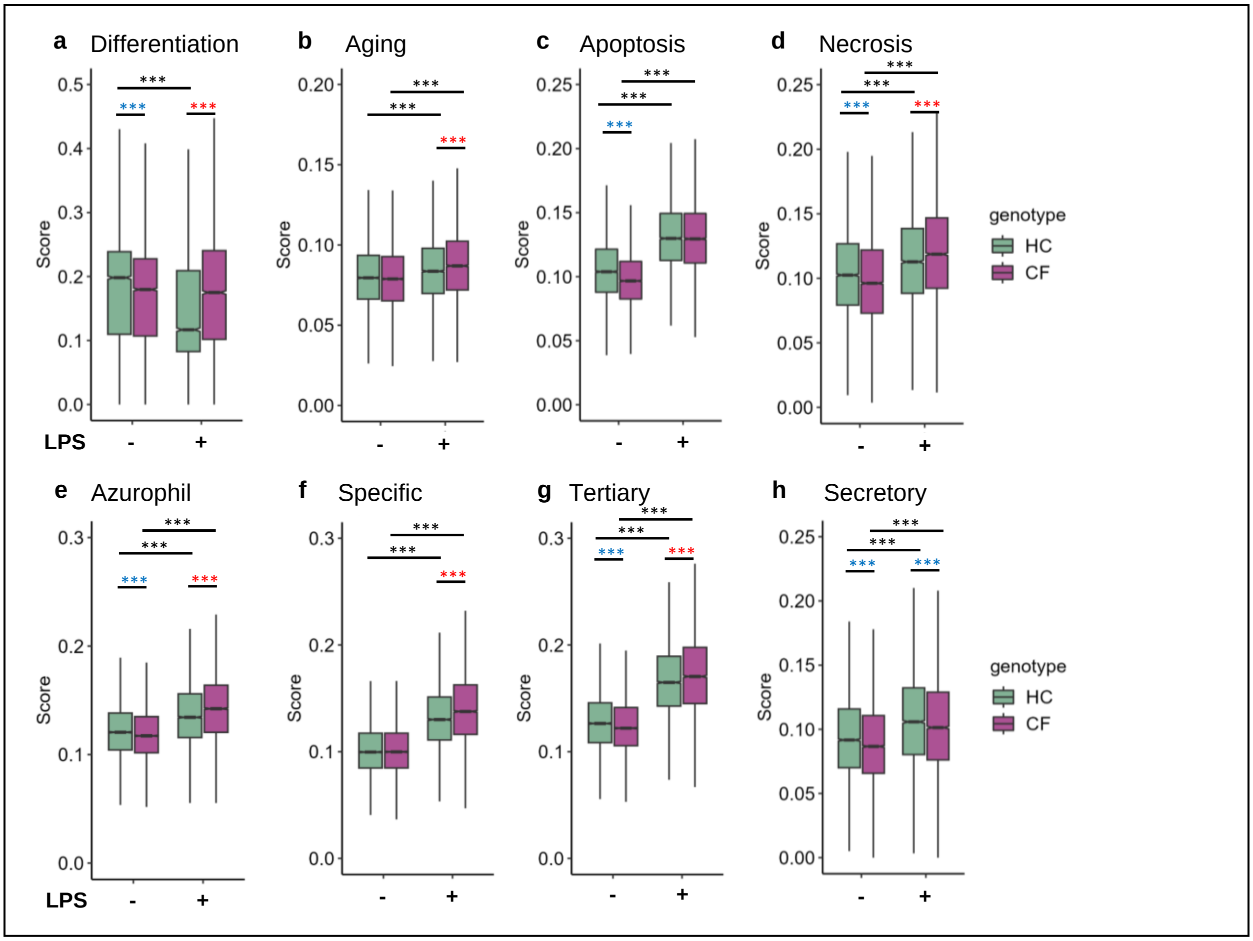


**Supplementary Fig. 7. Property scores of HC and CF neutrophils in basal state and upon LPS-stimulation. a-d**, Differentiation and aging scores; **e-h**, granule formation scores. Two-way ANOVA test was used to judge statistical significance between two comparing groups. *p< 0.05, **p< 0.01, ***p< 0.001. Red asterisks indicate CF neutrophils have significantly higher score, and blue asterisks significantly lower score than HC neutrophils in each comparison.
